# Impact of Lactobacillus GG on weight loss in post-bariatric surgery patients: a randomized, double-blind clinical trial

**DOI:** 10.1101/2024.03.24.24304808

**Authors:** Myra Nasir, Samuel Stone, Ian Mahoney, Justin Chang, Julie Kim, Sajani Shah, Laura A. McDermott, Paola Sebastiani, Hocine Tighiouart, David R. Snydman, Shira Doron

**Affiliations:** Department of Medicine, Division of Geographic Medicine and Infectious Diseases, Tufts Medical Center, Tufts University School of Medicine, Boston, MA, USA; Department of Medicine, Division of Cardiology, Tufts Medical Center, Boston, MA, USA; Department of Medicine, Division of Pulmonary Critical Care and Sleep Medicine, New York University, New York, NY, USA; Department of Surgery, Division of General Surgery, University of Illinois Chicago, Chicago, IL, USA; Department of Surgery, Mount Auburn Hospital, Cambridge, MA, USA; Department of Surgery, Division of Bariatric Surgery, Tufts Medical Center, Boston, MA, USA; Department of Medicine, Institute for Clinical Research and Health Policy Studies, Tufts Medical Center, Tufts University School of Medicine, Boston, MA, USA; Department of Medicine, Tufts Clinical and Translational Science Institute, Tufts University, Boston, MA, USA

## Abstract

**Introduction and Objectives:** There is increasing evidence suggesting the impact of human gut microbiota on digestion and metabolism. It is hypothesized that the microbiome in obese subjects is more efficient than that in lean subjects in absorbing energy from food, thus predisposing to weight gain. A transformation in gut microbiota has been demonstrated in patients who have undergone bariatric surgery which has been positively associated with post-surgical weight loss. However, there is lack of studies investigating the impact of probiotics on weight loss in post-bariatric surgery patients. The objectives of our study were to investigate the impact of a probiotic, Lactobacillus GG (LGG), on weight loss and quality of life in patients who have undergone bariatric surgery.

**Methods:** The study was registered with ClinicalTrials.gov NCT01870544. Subjects were randomized to receive either LGG or placebo capsules. Percent total weight loss at their post-operative visits was calculated and differences between the two groups were tested using a t-test with unequal variances. The effect of LGG on Gastrointestinal Quality of Life Index (GIQLI) scores was estimated using a mixed model repeated measures model.

**Results:** The mean rate of change in percent total weight loss at the ‘30-day’ post-operative visit for the placebo and treatment groups was 0.098 and 0.079 (p = 0.41), respectively, whereas that at the ‘90-day’ post-operative visit was 0.148 and 0.126 (p = 0.18), respectively. The difference in GIQLI scores on ‘30-day’ and ‘90-day’ visits were 0.5 (−7.1, 8.0), p=0.91 and 3.7 (−4.9, 12.3), p=0.42, respectively. LGG was recovered from the stools of 3 out of 5 subjects in the treatment group.

**Conclusion:** We did not appreciate a significant difference in the mean rate of weight loss or GIQLI scores between the groups who received LGG versus placebo. This study demonstrated survival of lactobacillus during transit through the gastrointestinal tract.

## Introduction

Obesity is a major public health problem worldwide. In the United States, 66% of the population is classified as overweight or obese [1].

The ‘microbiome revolution’ has, in the last 20 years, brought our attention to the importance of human gut microbiota as a factor affecting health and disease [2]. Gut microbiota include a range of microorganisms that inhabit the gastrointestinal tract and have been implicated in digestion and metabolism, protection from pathogens, insulin resistance and neurologic functioning [3-7]. Metagenomics and twin studies have revealed that certain microbial compositions are associated with lean body composition and alteration of these microbial communities is associated with weight gain and metabolic disease [8,9]. In obese subjects, the flora of the gut has been found to be less diverse, less rich and to have an increased ratio of Firmicutes to Bacteroidetes as compared to normal weight individuals [10]. It is hypothesized that the obese microbiome is more efficient than the lean microbiome in absorbing energy from food, thus predisposing to weight gain [8].

Bariatric surgery is the most effective treatment for the achievement of prolonged weight loss and has been associated with improvement in diabetes and hypertension [11-12]. Studies suggest that the beneficial health effects of these surgeries are not only explained by procedure-induced food restriction and malabsorption but also by alterations of neuroendocrine and immune signaling pathways [13]. An increase in Bacteroidetes and Proteobacteria with a decrease in Firmicutes after bariatric surgery has been demonstrated. When the Bacteroidetes/Firmicutes ratio after bariatric procedures was used as surrogate marker for the observed changes in the obese microbiome compared to the lean microbiome, it was positively associated with post-surgical weight loss [14].

Lactobacillus strain GG (LGG) is a widely studied probiotic, especially with regards to its influence on metabolic syndrome. A favorable impact on obesity and diabetes has been demonstrated in mice with diet-induced obesity showing evidence of improvement in insulin resistance with LGG [15,16].

Whereas there is increasing evidence to suggest that use of probiotics is linked with lower inflammatory state and weight loss, there is a lack of studies investigating the impact of probiotics on weight loss in patients who have undergone bariatric surgery [17-19].

We conducted a pilot trial to investigate the impact of LGG on weight loss in patients who have undergone bariatric surgery.

## Methods

This was a randomized, double-blind placebo-controlled pilot trial to investigate whether weight loss and changes in quality of life differ after bariatric surgery in patients who did and did not receive LGG. The primary outcome is the rate of change of percent total weight loss. Secondary outcomes are Gastrointestinal Quality of Life Index (GIQLI) at day of surgery, at 30-day visit and at 90-day visit, adverse event percentages and recovery of LGG from stool.

### Study population

Subjects were recruited from Tufts Medical Center in Boston, Massachusetts, USA, from June 24, 2013 to January 31, 2016, if they were scheduled for elective gastric surgery for weight reduction. Patients were eligible for the study if they were at least 18 years of age, were evaluated for surgery as outpatients, and had stable comorbidities. Exclusion criteria included active colitis; recent or planned receipt of radiation or chemotherapy; being on active immunosuppressive medication; known or suspected allergies to probiotics, lactobacillus, milk protein, or microcrystalline cellulose, or allergy to 2 or more of the rescue antibiotics that might be used to treat Lactobacillus infection (ampicillin, clindamycin and moxifloxacin) should that become necessary; and positive baseline stool culture for LGG.

Subjects included in the study were restricted from consuming certain yogurts, however since yogurt is a dietary staple after bariatric surgery, to improve enrollment after initial slow accrual, the protocol was later amended to allow for types of yogurt that contained select live cultures that would be expected to have minimal effect on patients’ microbiota, as well as yogurt without cultures. The four commercially available yogurts that were permitted were Yoplait Greek, Breyer’s light, Brown Cow Fat Free Greek and Brown Cow Greek.

Study patients undergoing sleeve gastrectomy were initially planned to be excluded from the study. However, sleeve gastrectomy rapidly became the preferred weight-loss surgery which resulted in significantly fewer gastric bypass surgeries at Tufts Medical Center than they were at the time of initial study design. Therefore, in order to improve enrollment, the study design was amended to included patients undergoing sleeve gastrectomy, in addition to gastric bypass surgery.

### Trial Design and Oversight

The trial design is parallel with an allocation ratio of 1:1. Sample size analysis was not conducted since this was a pilot study to inform a future larger scale trial.

The random allocation was done by the statistician using a random number generator with the allocation sequence provided directly to the research pharmacy who labeled the pills with the subject names but without identification of the compound. Study staff, which included investigators and research assistants, enrolled subjects and went to the research pharmacy to pick up the study compound already labeled for the subject. Thus, investigators, study staff and participants were blinded, as were the clinical staff caring for these subjects.

Subjects were randomized to receive either probiotic capsules of LGG (1x10^10^ organisms per capsule) or identical appearing placebo capsules provided by Martek, manufactured by Chr Hansen, Inc., to be taken twice daily for 44 days, beginning 14 days before surgery. Pill bottles contained 100 capsules. Capsules were collected from each subject prior to initiation of the course and at study completion. One capsule from each time period was cultured to confirm viability of the organism and organism counts. Ten extra capsules were provided in case of loss of capsules. Remaining capsules were counted at the end of the study period to assess adherence. Stool samples were obtained at baseline (2-4 weeks before surgery), on the day of surgery and at ‘30-day’ and ‘90-day’ post-operative visits to assess for the presence of colonization by LGG.

To monitor for adverse events, subjects completed questionnaires about the presence or absence of certain symptoms before and after they began taking the study compound. These questionnaires were completed at the first surgeon visit, first pre-operative visit, and day of surgery in order to establish a baseline, then at the first four weekly post-operative telephone check-ins, ‘30-day’ post-operative visit and ‘90-day’ post-operative visit.

All subjects received routine pre-operative prophylactic antibiotics to prevent surgical site infection. One subject in the placebo group was receiving azithromycin pre-operatively for suspected bacterial infection and continued it for several days after surgery.

The study was registered with ClinicalTrials.gov NCT01870544, and was approved by the Tufts Medical Center Institutional Review Board. A written informed consent was taken from all subjects. CONSORT reporting guidelines were used (S1 CONSORT Checklist) [20].

### Data collection

Demographic information, history of chronic diseases, and medications were collected at screening. Participants were asked to complete the validated GIQLI questionnaire at the pre-operative visit, on the day of surgery, and at the post-operative visits closest to 30 and 90 days after surgery [21]. Study weight measurements were taken at the preoperative visit, the day of surgery, and at the visits closest to 30 and 90 days after surgery. Since these visits did not necessarily occur on post-op days 30 and 90, we refer to them throughout this manuscript as the ‘30-day’ and ‘90-day’ visits in quotation marks. Weights were also extracted from the electronic medical record when available from visits not part of the study.

### Statistical analysis of clinical outcomes

Compared to other metrics such as excess weight loss and change in body mass index, percent total weight loss (%TWL) has been found to have the least variability when stratified by preoperative patient characteristics, and therefore is the metric used in this study [22]. Subject weights were measured during post-operative visits. Percent total weight loss at X days (%TWL_X) was calculated as (weight at surgery – weight at visit)/ weight at surgery x X/(delta day), where delta day was the difference in days between the date of surgery and the date of visit. Differences in %TWL_X between the two groups were tested using a t-test with unequal variances.

### Sample collection

Subjects were given stool collection kits consisting of a toilet hat, gloves, tongue depressors, specimen cups and paper bags and asked to produce and save a stool sample in their freezer within 24 hours of their appointment date and to bring the sample to their scheduled visit. Samples were then aliquoted and frozen at -80°C.

### Isolation of LGG from Capsule and Stool samples

Stool samples and capsules were serially diluted and plated in duplicate onto Lactobacilli selective agar (BBL) supplemented with tomato juice and glacial acetic acid. Plates were incubated for 48 hours in an anaerobic chamber (5%CO_2_, 10%H_2_, 85% N_2_) at 35 ^+^/- 2° C and were then observed for characteristic growth and colonial morphology. Presumptive LGG colonies, appearing as white and creamy with a distinct buttery smell, were enumerated. The palisading appearance of LGG colonies on Gram stain was used to distinguish it from other Lactobacillus species. All strains presumptively identified were run through the APIZYM® (Biomerieux), an identification kit that differentiates between the Lactobacillus species on the basis of enzymatic reactions. Isolates that were consistent with LGG by APIZYM® were additionally run through the API CH-50 ® (Biomerieux) system, which differentiates between different species of Lactobacillus on the basis of carbohydrate fermentation.

### Statistical analysis of Gastrointestinal Quality of Life Index (GIQLI) scores

The effect of LGG on the continuous GIQLI scores was estimated using a mixed model repeated measures model. The outcome for each subject consisted of all follow-up measurements including the baseline measurement. All variables included in the model were treated as fixed effects and included categorical time, randomization and their interaction. We performed sensitivity analyses to account for missing data in follow-up using pattern mixture models (PMM). We used the core functionality available in SAS PROC MI and MIANALYZE procedures to implement two types of PMM: standard PMM and control-based PMM.

## Results

A total of 37 bariatric surgery candidates were recruited and followed up through April 2016. Of these, 15 subjects withdrew prior to randomization, 3 after randomization and before study drug was administered and 1 who received study drug but was lost to follow up, with 18 subjects completing follow-up (Figure 1). Ten subjects received oral placebo and 8 received oral LGG. Table 1 compares the characteristics of subjects by study group. The median age of the placebo group and the treatment group was 47.5 and 46.5, respectively. Out of 18 subjects, 12 were female, comprising 60% of the placebo group and 75% of the treatment group. Of those who received LGG, compliance was variable and ranged between 0 and 19 doses missed. Baseline median body mass index (BMI) and excess weight of the placebo group (42.44 and 13.02 lbs, respectively) were comparable to the baseline median BMI and excess weight of the treatment group (41.16 and 102.28 lbs). The placebo group, as compared to the treatment group, had a higher percentage of patients with diabetes (40% vs. 25%, respectively) and a lower percentage of patients with hyperlipidemia (20% vs. 25%, respectively). Most subjects underwent sleeve gastrectomy (70% and 75% for placebo and treatment groups, respectively) while the rest underwent roux-en-y gastric bypass.

**Table 1:** Characteristics of the study population.

|  | N = 10 | N = 8 |
| --- | --- | --- |
| Characteristic | Placebo | Treatment |
| Age, median (IQR), y | 47.5 (40.75 – 49.75) | 46.5 (40.0 – 53.0) |
| Female, No. (%) | 6 (60%) | 6 (75%) |
| BMI, median (IQR) | 42.44 (41.52 – 45.80) | 41.16 (40.14 – 42.96) |
| Diabetes, No. (%) | 4 (40%) | 2 (25%) |
| Hyperlipidemia, No. (%) | 2 (20%) | 2 (25%) |
| Excess Weight, median (IQR), lbs. | 103.02 (99.79 – 127.13) | 102.28 (86.60 – 120.54) |
| Quality of Life score, mean (SD) | 59.0 (6.1) | 59.4 (7.2) |
| Surgery Type |  |  |
| Roux-en-y gastric bypass, No. (%) | 3 (30%) | 2 (25%) |
| Sleeve gastrectomy, No. (%) | 7 (70%) | 6 (75%) |

**Figure 1.**
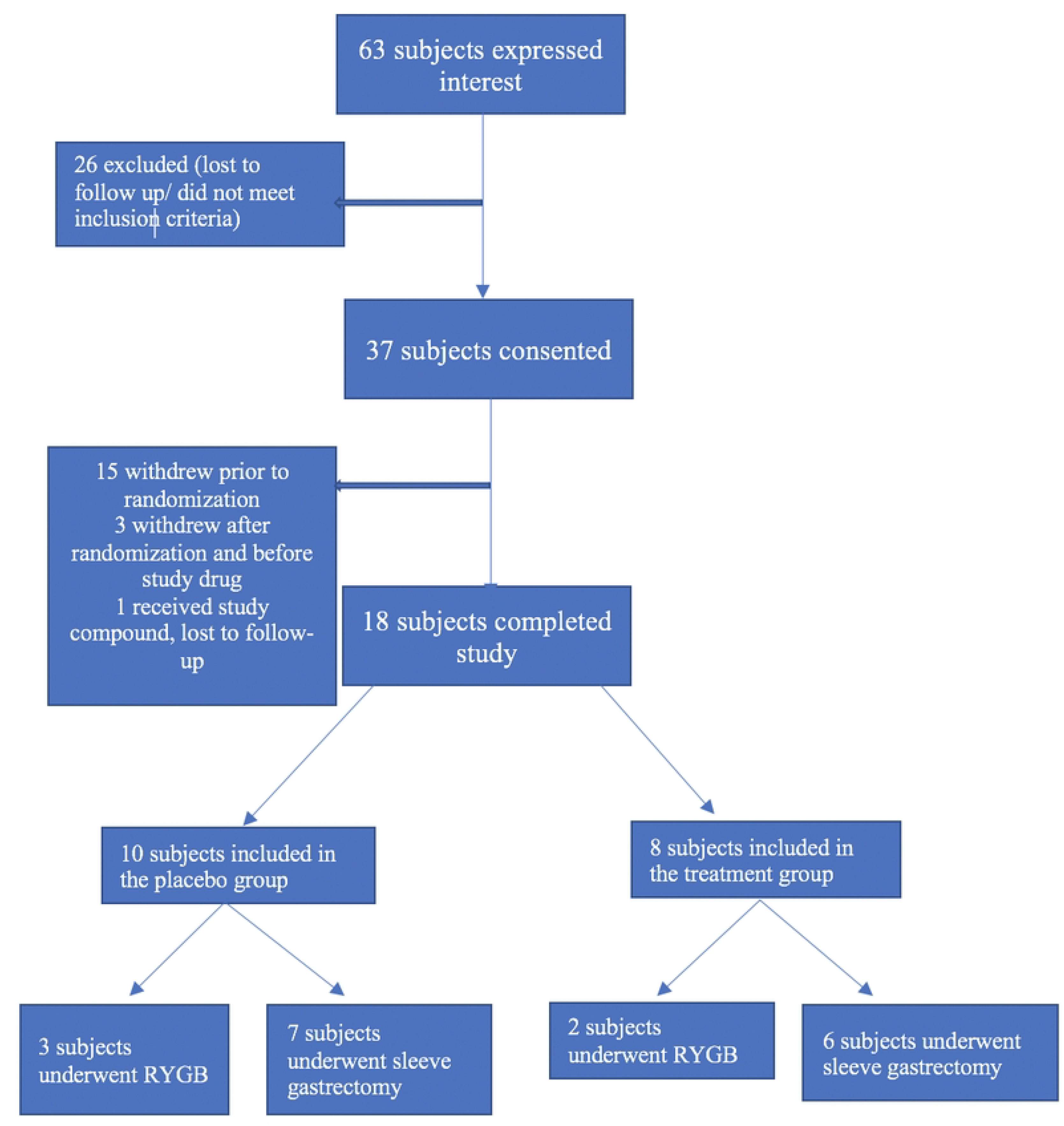
Flowchart of subject selection RYGB: Roux-en-y gastric bypass

At baseline, mean (SD) GIQLI scores for treatment and placebo groups were 59.0 (6.1) and 59.4 (7.2) respectively. On the day of surgery, GIQLI score was 61.1 (6.3) for the treatment group as compared to 57.4 (9.2) for the placebo group with a mean difference (95%CI) of 3.7 (−3.8, 11.2), p=0.35. At the ‘30-day’ post-operative visit, GIQLI score was 55.1 (8.5) for the treatment group as compared to 55.5 (4.0) for the placebo group with a difference of 0.5 (−7.1, 8.0), p=0.91. At the ‘90-day’ post-operative visit GIQLI score was 60.7 (4.9) for the treatment group as compared to 56.5 (6.8) for the placebo group with a difference of 3.7 (−4.9, 12.3), p=0.42.

The mean time between surgery and the ‘30-day’ post-operative visit for the treatment and placebo groups was 34 and 54 days, with a median of 38 and 48 days, respectively. The mean time between surgery and the ‘90-day’ post-operative visit for the treatment and placebo groups was 105 and 125 days, with a median of 107 and 111 days, respectively. The mean rate of change in percent of total weight loss (%TWL) at the ‘30-day’ post-operative visit for the placebo and treatment groups was 0.098 and 0.079 (p = 0.41), respectively, whereas that at the ‘90-day’ post-operative visit was 0.148 and 0.126 (p = 0.18), respectively.

LGG was not recovered from any stool samples from subjects belonging to the placebo group. Among subjects in the treatment group, no samples were successfully obtained after completion of the course of treatment. All samples were taken during the treatment course. Stool samples were collected from only 5 of the 8 subjects in the treatment group after the start of the course of treatment. Of the 5, LGG was recovered from the stool samples of 3 subjects. One of the subjects from whom LGG was not recovered was later found to have missed 19 doses, and the other 4 doses.

The most common adverse events experienced by the study and placebo groups while taking the study compound were increased frequency of bowel movements, loss in appetite, flatulence, constipation, rumbling, looser stools, and abdominal fullness (Table 22).

**Table 22:** Adverse events reported by subjects while taking study compound.

| Adverse events | Study group | Placebo group |
| --- | --- | --- |
| Increased frequency of bowel movements | 6 | 8 |
| Loss in appetite | 6 | 7 |
| Flatulence | 6 | 7 |
| Constipation | 7 | 5 |
| Rumbling | 3 | 7 |
| Looser stools | 5 | 6 |
| Abdominal fullness | 5 | 6 |
| Burping/belching | 4 | 6 |
| Abdominal pain | 4 | 2 |
| Bloating | 3 | 2 |
| Nausea | 3 | 2 |
| Urgent bowel movements | 2 | 3 |
| Trouble swallowing food | 1 | 3 |
| Other illnesses | 3 | 1 |
| Heartburn | 0 | 3 |
| Fevers/chills | 1 | 2 |
| Blood in stool | 2 | 1 |
| Hospitalization | 2 | 0 |
| Increased gas | 1 | 1 |
| Fluid coming up in the mouth | 0 | 1 |

## Discussion

Growing evidence supports that weight-loss interventions, whether dietary or surgical, lead to changes in the composition of the gastrointestinal microbiome [23]. However, there are conflicting data regarding the impact of probiotics on weight loss. Whereas Kadooka et al demonstrated a significant decrease in weight and body fat in subjects who received fermented milk with lactobacillus compared to a control group [24], a meta-analysis by Mohammadi et al of 9 randomized controlled trials including 410 subjects showed no significant difference in BMI or percentage of excess weight loss between the probiotic and control groups [25].

In our pilot study of patients who underwent bariatric surgery, we did not appreciate a clinical or statistically significant difference in the mean rate of weight loss between the groups who received LGG versus placebo. Our study findings are in keeping with results demonstrated by a systematic review and meta-analysis of 5 randomized controlled trials including a total of 279 patients after bariatric surgery in which no significant difference in percent excess weight loss at 3 months, change in BMI, waist circumference change or fat mass change was noted between the probiotic and placebo groups [26]. However, it should be noted that if there were some minor impact of oral probiotics on weight loss, this may be masked by the substantial effect of bariatric surgery on weight loss, resulting in no measurable difference between the two groups.

This is the first study to our knowledge to evaluate whether probiotics can be recovered in the stool of patients who have undergone bariatric surgery. To exert its beneficial properties, probiotics must survive during their transit through the low pH environment of the stomach and lytic enzymes present in the small intestine. In our study, LGG was recovered from 3 of 5 subjects in the treatment group, demonstrating survival, though inconsistent, of the lactobacillus during transit through the gastrointestinal tract. Several factors may have contributed to the lack of recovery of LGG from 2 subjects in the treatment group who were still in the treatment phase, including adherence, the constituents of the subjects’ diet which can affect survival and growth of the bacteria, the stability of the culture and size of the inoculum which affect the survival of probiotic bacteria.

There was no significant difference in the GIQLI scores within the follow-up period, between the study and placebo groups.

In our pilot study of patients undergoing bariatric surgery, LGG was found to be safe. Adverse event rates were similar in the two groups, and consistent with what would be expected in patients who have undergone gastrointestinal surgery.

Our study had several limitations. It was a pilot and therefore not intended to be fully powered to detect statistically significant differences in outcomes between groups. Though the follow-up outpatient visits were intended to be at 1- and 3-months post-surgery, there was significant variability in the number of days after surgery at which subjects were evaluated. The 3-month duration of follow-up may not have been long enough to demonstrate a clinically significant difference in weight loss or quality of life scores between the two groups.

## Data Availability

All relevant data are within the manuscript and its Supporting Information files.

## Funding

This study was supported by a grant from Tufts University Collaborates and NIH CTSA grant UM1TR004383 and UM1TR004398 awarded to SD, JK and DS.

S1 CONSORT Checklist

S2 Appendix. Study Protocol

